# COVID-19 Origins: Quantifying Scientific Consensus Amid Political Polarization Through Mixed-Methods Meta-Analysis

**DOI:** 10.1101/2025.06.06.25328995

**Authors:** C. O Afolabi, J.D Adekunle, M. I Oyeniran, S. O Oyelakin, C. K Ogu, E. J Ayanlowo, C. O Robert, H. S Sule, G. E Ideh, S.A Alagbe, O.O Fagbemiro, Y. A Adeniyi, T. I Adegboyega, O.O Samsudeen, K. O Badru, K. O Shakioye, A. T Alimi, A. A Amos, B. N Ebonyem

## Abstract

From 2019 till now, the origin of COVID-19 remains a debate between scientists and politician specially between the US and China. Majorly, two independent hypotheses were postulated. The lab leak hypothesis, which involves an escape of the virus from WIV laboratory in Wuhan, China and the natural origin hypothesis possible through an intermediate host. These hypotheses had been discussed since the pandemic with no definite conclusion. However, there is a belief of what the origin might be within the science community that might not be influenced politically. We aimed to check the direction of scientific consensus on the origin matter and to discuss the effect and impact of politicization. To achieve this, a mixedmethod meta-analysis involving a content-based qualitative and quantitative synthesis was conducted. 48 studies were selected using a PRISMA model and were synthesized on MASQDA. The protocol was registered on PROSPERO with an ID: 1055566. The result showed that majority of the scientist support the natural origin of covid 19 and the sentiment around this hypothesis is positive (0.398), indicating a more optimistic and affirmative language compare to the lab leak hypothesis with a negative average sentiment score of −0.124, suggesting that discourse around this theory is comparatively more negative. The lab-leak hypothesis fail due to the following: (1) Lack of Genetic Evidence for Engineering, (2) No Pre-Existing Virus Matching SARS-CoV-2 in Labs, (3) Furin Cleavage Site (FCS) Is Naturally Occurring and experimental attempts to generate an FCS in bat coronaviruses failed, suggesting natural evolution, (4) Early Cases Linked to Animal Exposure, Not Labs, (5) Historical Precedent for Natural Zoonotic Spillover: SARS-CoV-1 (2003) and MERS-CoV (2012), (6) Lack of Credible Evidence for Lab Involvement: No scientific publication, leaked document, or whistleblower testimony

## INTRODUCTION

The COVID-19 pandemic, caused by the novel severe acute respiratory syndrome coronavirus 2 (SARS-CoV-2) as named by the ICTV on 11 February 2020, has emerged as one of the most significant global crises of the 21st century [1][2]. Since its first reported cases in Wuhan, China, in late 2019, the pandemic has claimed millions of lives and upended socioeconomic systems worldwide [3][2]. While much scientific attention has been devoted to understanding its transmission [4], treatment [5][6], and prevention, one of the most enduring and controversial questions remains: What is the true origin of SARS-CoV-2?

The debate over the origin of COVID-19 has largely revolved around two mutually exclusive primary hypotheses: (1) a zoonotic origin, involving a natural spillover from an animal host to humans, and (2) a laboratory-related origin, potentially involving an accidental release from Wuhan Institute of Virology (WIV) research facility in China [2][7]. These two hypotheses are not merely academic or scientific; they touch upon sensitive geopolitical dynamics, national accountability, and the integrity of global scientific collaboration. In 2021, the World Health Organization (WHO) early investigative mission to China on COVID-19 issues stated that its “extremely unlikely” the origin of COVID-19 was as a result of a lab leak, but later acknowledged the need for further investigation into all plausible hypotheses. Meanwhile, multiple governments, intelligence agencies, and independent scientists have released conflicting statements, further introducing more complexity. Public discourse has also been shaped by misinformation, ideological biases, and politically motivated narratives. Indeed, what began as a virological inquiry has gradually escalated into a battleground of media sensationalism, political posturing, and social polarization.

However, understanding the origin of SARS-CoV-2 is not just an exercise in historical curiosity. It centers on informing future pandemic preparedness, shaping biosafety protocols, and guiding public health policies [8][9]. It is equally important to examine how origin narratives are constructed, communicated, and politicized [7][10]. These narratives significantly influence public perception, government responses, and scientific inquiry [10]. They also affect international diplomacy, particularly between major global powers such as the United States and China, whose relationship has been tested by accusations and counter-accusations regarding COVID-19s genesis [10][7].

Therefore, investigating origin narratives must go beyond identifying the biological starting point of the virus. It must also explore how scientific evidence, political discourse, and media representations interact to shape the global understanding of COVID-19s origins. This layered analysis can offer critical insights into the intersection of science, politics, and society in the 21st century. In light of the complex and multifaceted nature of the COVID-19 origin debate, this study seeks to explore the issue from an interdisciplinary perspective, employing both qualitative and quantitative methods with the aim of looking at the dominant narratives surrounding the origin of COVID-19 and how they have evolved over time. The scientific evidence supports each major hypothesis (zoonotic vs. lab-based), and how has this evidence been represented in academic and public discourse, and to what extent has the investigation into COVID-19’s origins been influenced by political, institutional, and geopolitical factors? Finally, to check how the politicization of the origin debate has affected scientific communication, research integrity, and public trust.

### INTERMEDIATE HOST

Scientifically, a host is an animal or plant on or in which a parasite or commensal organism lives[11]. According to the Biology Online Dictionary, there are five types of hosts, which are: primary, secondary, paratenic, accidental, and reservoir [12]. Out of these, the role of secondary host (also known as intermediate host) in diseases and pathogen transmission is significant. The definition of secondary host comes from its ability to serve as a passage(i.e., intermediate) for pathogen transmission. An intermediate host is an organism that temporarily harbors a pathogen, such as a virus, allowing it to replicate or mutate before the pathogen is transmitted to its final or primary host, often a human[14][13][15]. In zoonotic diseases (infections that jump from animals to humans), the intermediate host acts as a biological bridge between the natural reservoir and humans[16][17]. The primary role of an intermediate host is to facilitate viral adaptation and amplification[18][19]. Within the intermediate host, the virus may increase in concentration and undergo genetic changes or recombination, which can enhance its ability to bind to human receptors and cause infection[20][21][64]. While animals are overwhelmingly the known intermediate hosts in zoonotic transmission, theoretically, humans can also act as intermediate hosts in anthroponotic or reverse-zoonotic eventswhere a virus originates in animals[17], infects humans, and is then passed on to other animals or other humans with evolutionary shifts[22][23][24][25]. However, in classical zoonotic emergence (such as SARS, MERS, and potentially COVID-19), intermediate hosts have always been animals(Figure 1).

**Figure 1:**
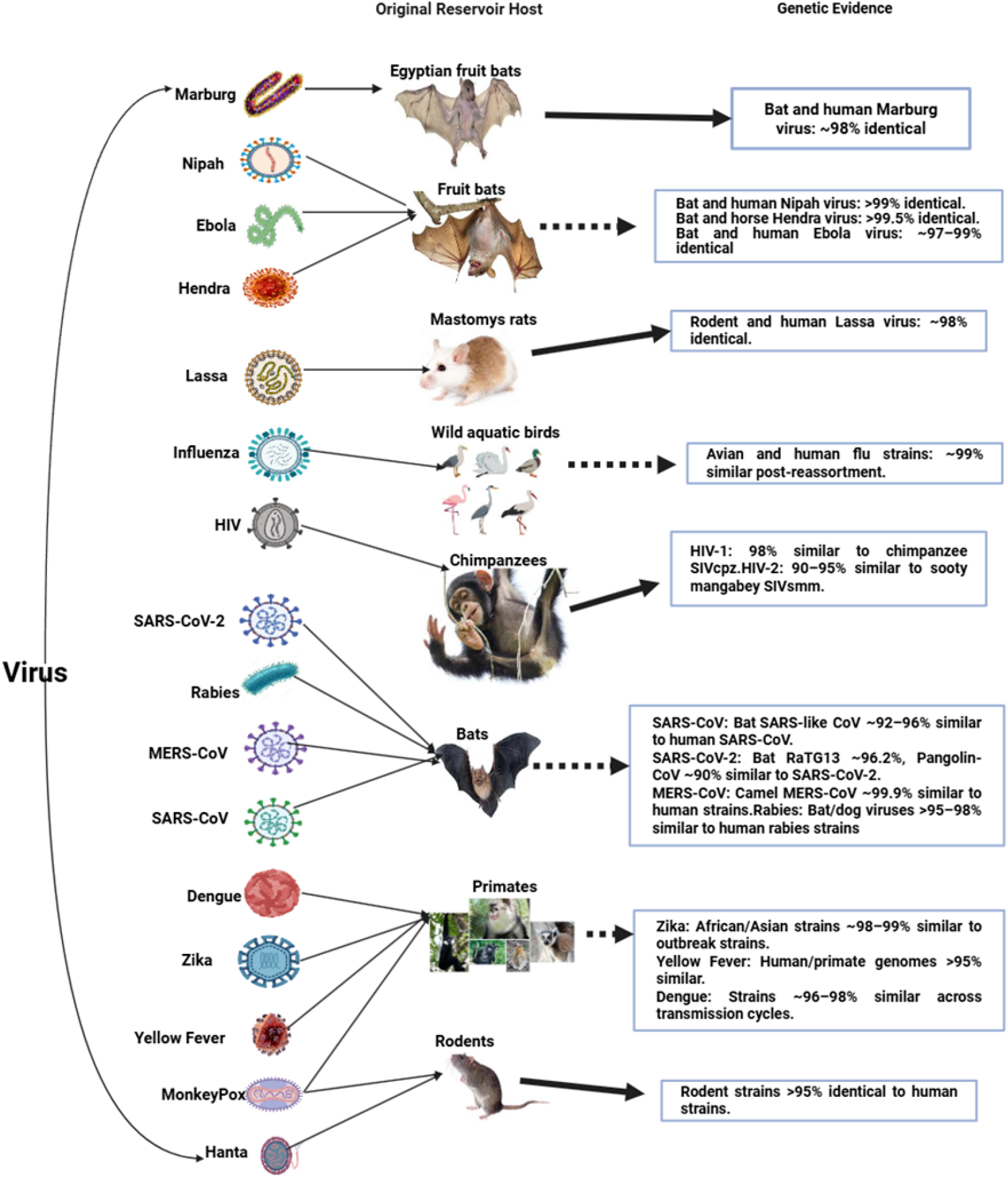
The thick dotted lines indicate an intermediate host, while the thick line indicates a direct host. Viruses such as HIV (1 & 2) [26], Marburg virus [27], Rabies virus [28], Hantavirus [29], Monkeypox virus (Mpox) [30], and Lassa virus [31] directly infected the human population. Dengue virus, Yellow Fever virus, and Zika virus were transmitted by Mosquitoes (Aedes spp.) [32][33][34]. Influenza A and Nipah virus was intermediated by Pigs, poultry [35][36], Hendra virus intermediated by Horses [37], Ebola virus Possibly by primates [38], MERS-CoV intermediated by Dromedary camels [39], SARS-CoV intermediated by Civet cats [40], while SARS-CoV-2 possibly by pangolins [41].

## METHODOLOGY

### Research design

Given the interdisciplinary and contentious nature of the COVID-19 origin debate, a single-method study would fail to capture the full complexity of the issue. To avoid this, we conducted a mixed-method meta-analysis. This method involves the extraction and synthesizing of qualitative and quantitative data from findings across multiple sources and disciplines [42]. In the context of COVID-19, this synthesis is critical because no single study or perspective has definitively resolved the origin debate while the Meta-analysis allows for the integration of diverse viewpoints, identification of consensus (or lack thereof), and assessment of methodological strengths and weaknesses across studies [43]. The quantitative component of this study involves systematic data aggregation and statistical synthesis from peer-reviewed literature and public databases [42]. Metrics such as the frequency of specific origin-related claims, evolution of scientific consensus over time, co-authorship networks across geopolitical boundaries, and citation impact were quantitatively analyzed. Similarly, the qualitative method was used to explain why certain narratives gain traction or how political and ideological forces shape scientific inquiry through thematic content analysis [42][44].

### Search strategy

We focused on peer-reviewed articles, government reports, and official statements published in PubMed, Google Scholar, SAGE Journals, and the official website of the World Health Organization (WHO). These databases were queried using a combination of predefined keywords and Boolean operators, including: (COVID-19 OR SARS-CoV-2) AND (origin OR source OR spillover OR zoonotic OR lab leak OR Wuhan OR animal origin). Additional keyword phrases were used to enhance search sensitivity, such as COVID-19 origin, SARS-CoV-2 origin, origin of COVID-19, and origin of SARS-CoV-2. Studies were selected based on predefined inclusion and exclusion criteria. Inclusion criteria were: (1) the study focused specifically on the origin of COVID-19, (2) it was published between 2019 and 2025, (3) it was written in English, and (4) it was available as open-access full-text. Eligible study types included peerreviewed journal articles, systematic reviews, meta-analyses, narrative reviews, and institutional or governmental reports. Studies were excluded if they (1) did not focus on the origin of COVID-19, (2) were published before 2019, (3) were written in a language other than English, or (4) were not available as open access. Editorials, commentaries, and letters without substantial analytical content were also excluded. The review protocol was registered with PROSPERO (ID: 1055566). The initial search with the keywords returned a total of 142,314 records, including 5,286 from PubMed, 53 from SAGE Journals, approximately 124,000 from Google Scholar (top 500 screened by relevance), and 50 from Elsevier (Figure 2).

**Figure 2:**
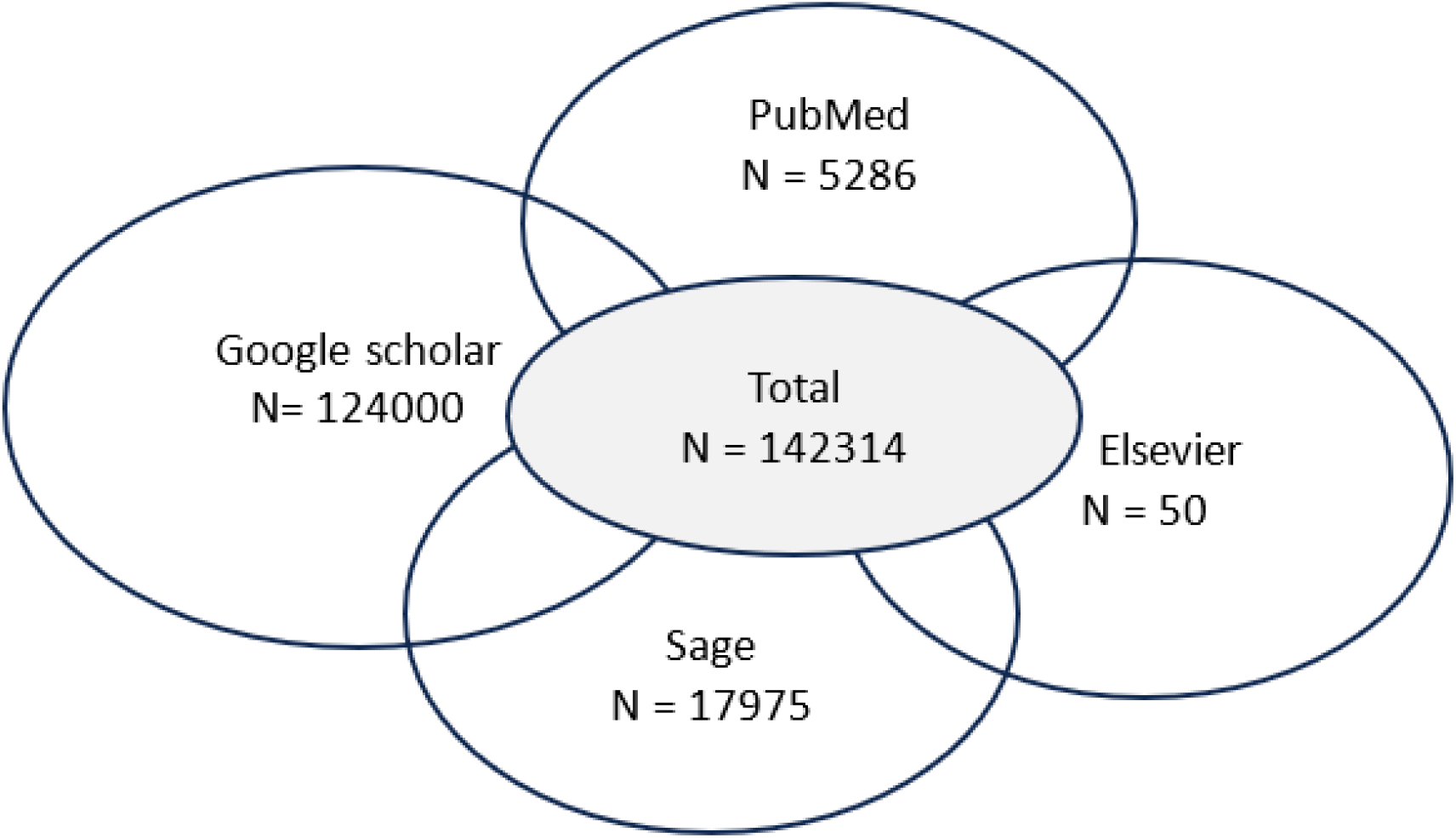
Number of identified studies across databases

After the first search, 2,319 duplicates were removed and 145,000 studies were screened. A total of 144,100 records were excluded after which 900 studies were sought for retrieval and 50 were not retrieved. On applying the aforementioned inclusion and exclusion criteria, 628 studies were excluded. 222 studies were included in the review and 48 studies were synthesized (Figure 3) [Supplementary 1].

**Figure 3:**
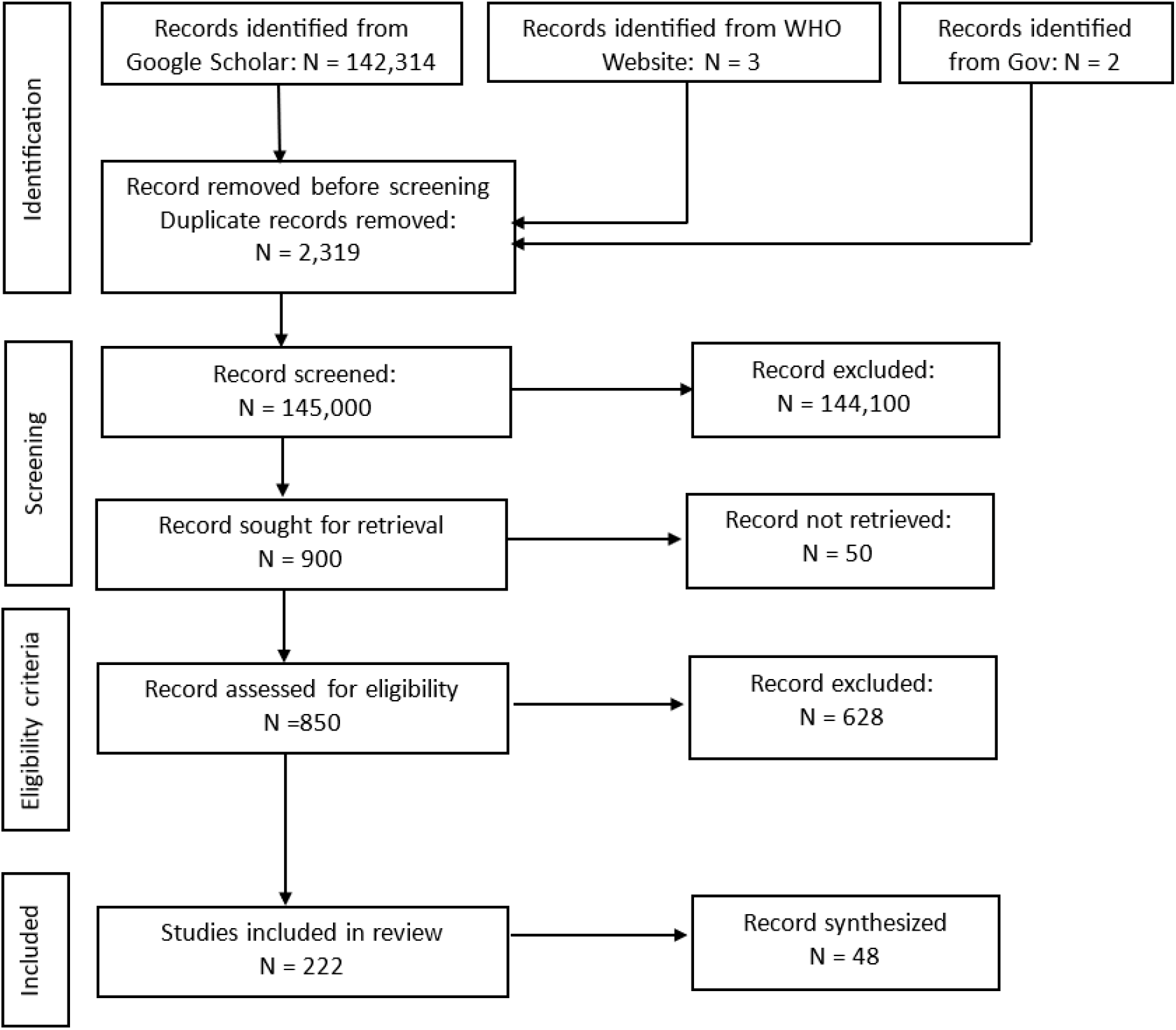
The Preferred Reporting Items for Systematic Reviews and Meta-Analyses (PRISMA) model indicating Study Selection Process [Supplementary 2]

### Qualitative Analysis

The thematic analysis was done to extract meaningful patterns, statements, and narratives from textual materials. Each of the identified documents was renamed with a Unique document ID (D1, D2, D3, Dn) for consistency and easy identification [Supplementary 3]. These IDs correspond to the study ID on the paper meta-data collected on an Ms. Excel sheet which contains: reference, title, Source, abstract, etc., and to the paper ID on the author meta-data collected on an Ms. Excel sheet containing author name, author field, author title, country, etc. [Supplementary 4]. The document (i.e., articles, reports, etc.) was carefully read multiple times to gain a general sense of recurring ideas and themes after which the text were broken into segments and manually codes for significant content. Codes were clustered into thematic categories based on conceptual similarity and frequency of appearance. The coding framework was done inductively and deductivelyinitial codes emerged from the literature, while others were based on theoretical frameworks such as framing theory and science-politics interface theory [45]. Themes were categorized into: Natural origin theory, lab-leak origin theory, scientific consensus, politicization of the COVID-19 origin, why lab-leak fails, prior event, Unknown origin, recommendation and future direction, and impact of global policy [Supplementary 5]. The coding was done on Maxqda qualitative analytic pro 2020 tool version 20.3.1[46].

### Quantitative analysis

The extracted data was standardized by tokenization, stop word removal, and lemmatization for comparison across sources. Descriptive statistics such as frequencies, proportions, means, medians, were done for temporal trend (annual publication/citation metrics) and source distribution. Hypothesis support was quantified through segment-level frequency analysis, comparing “Lab Leak” and “Natural Origin” prevalence across subgroups (author fields, institutions, countries) using a percentage breakdown. Textual analyses included TF-IDF-weighted keyword extraction, keyness metrics (likelihood ratio) [47], and sentiment scoring (AFINN lexicon) [48] to contrast lexical and affective features between hypotheses. Similarity metrics Jaccard [49][50][51] and Levenshtein[52][53] were used to mapped inter-document textual relationships, while targeted phrase extraction identified evidentiary keywords (e.g., “zoonotic,” “spillover) [Supplementary 6 & Supplementary 7]. Data were processed and analyzed using R statistical software.

### Validity, reliability, and limitations

Construct validity was ensured by clearly defining each thematic category and cross-verifying data sources. Internal validity was supported by triangulating findings from multiple sources and applying inter-coder agreement in qualitative coding. Similarly, external validity was cautiously considered, with an acknowledgment that findings may be more representative of English-speaking scientists. For qualitative data, inter-coder reliability was maintained by employing two researchers for the thematic coding process. A manual thematic analysis was deliberately done to avoid a semantic equivalence between a contradictive word (e.g., not originate naturally and originate naturally). For quantitative datasets, reliability was ensured by validating data collection procedures, running duplicate queries for verification, and using standardized meta-analytical methods. In this study, we acknowledged that the analysis primarily included English-language sources, potentially omitting important regional or non-Western perspectives resulting into a language bias. Some government reports and origin-related research may remain classified or inaccessible, introducing gaps. Given the evolving nature of COVID-19 origin debates, some trends and conclusions may shift with new evidence. Despite these limitations, the mixed-method meta-analytical design offers a comprehensive and balanced framework for understanding the COVID-19 origin debate and its broader socio-scientific implications [Supplementary 2]. %

## RESULTS AND DISCUSSION

Out of the 48 selected documents, majority of the papers were published in 2020(20[41.67%]) with 12,608(94.6%), 788.0 (s1273.8),80.5 (IQR: 0873.5) citations. This is likely due to the urgency and global focus on COVID-19 during the early phase of the pandemic. In contrast, papers from 2021 (15[31.25%]) and 2022 (6[12.50%]), 2023(5[10.42%]), 2024(2[4.17%], and 2025(1[2.08%]) experienced a significant decline in publication and citations.

**Table 1:** Distribution of included papers and citation metrics by year of publication.

| Year | Papers |  |  | Citations |  |  |
| --- | --- | --- | --- | --- | --- | --- |
|  | No. | % of Total | Total | Avg. (+SD) | Median (IQR) | % Zero |
| 2020 | 20 | 41.67 | 12 608 | 788.0 (+1273.8) | 80.5 (0–873.5) | 10.0% |
| 2021 | 15 | 31.25 | 476 | 43.3 (+51.2) | 14 (2–53) | 0.0% |
| 2022 | 6 | 12.50 | 199 | 33.2 (+56.1) | 11 (0–25.5) | 16.7% |
| 2023 | 5 | 10.42 | 41 | 10.3 (+19.2) | 1 (0–11.25) | 40.0% |
| 2024 | 2 | 4.17 | 2 | 1.0 (+0.0) | 1 (1–1) | 0.0% |
| 2025 | 1 | 2.08 | 0 | 0 (0) | 0 (0–0) | 0.0% |
| Total | 48 | 100.00 | 13 326 | – | – | 12.5% |

### The Origin Debate: Zoonotic hypothesis and Lab-origin hypothesis

A total of 699 discrete statements were identified that explicitly supported either the Lab Leak or the Natural Origin Hypothesis regarding the origin of COVID-19. There were 111(15.9%) statements that supported the Lab Leak Hypothesis and 588(84.1%) statements supported the Natural Origin Hypothesis (Figure 4).

**Figure 4:**
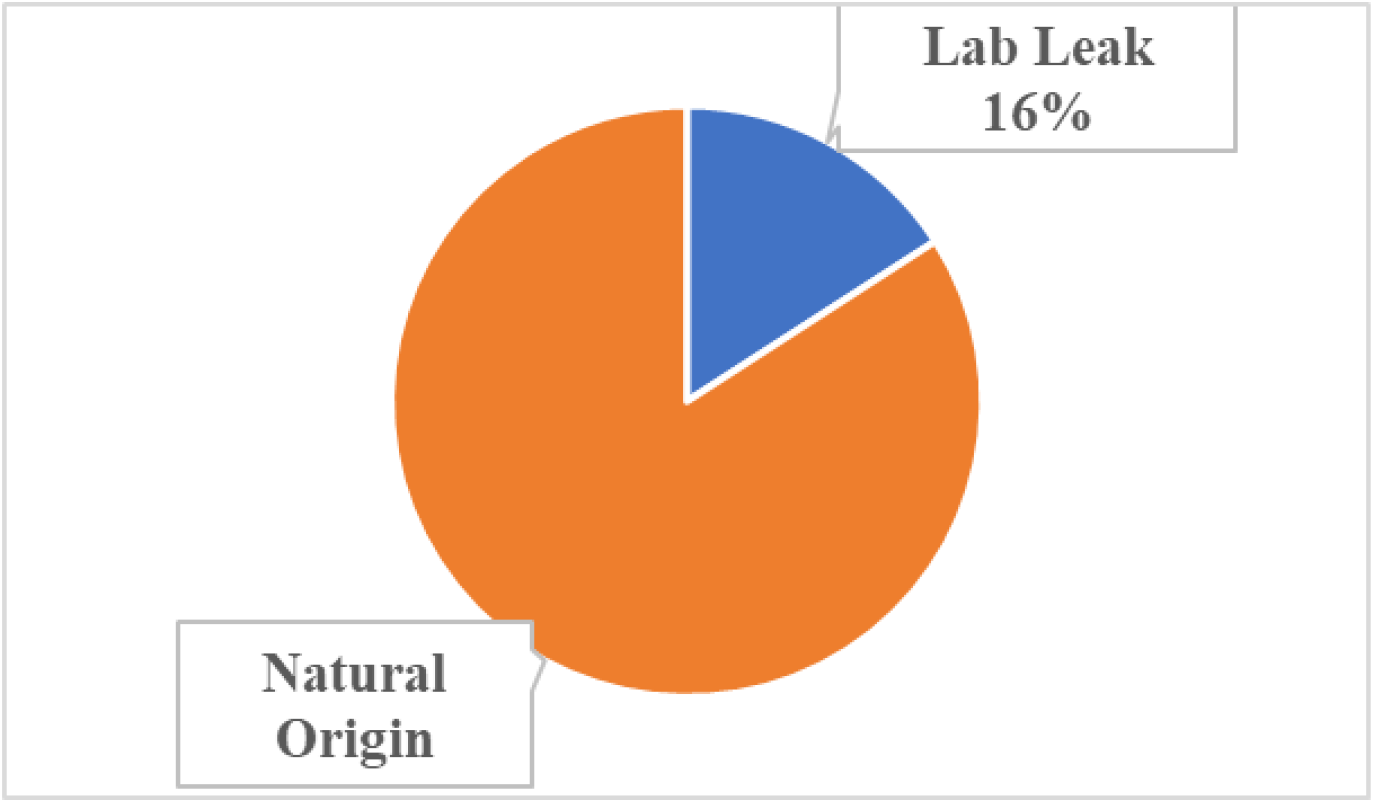
Distribution of discrete statements supporting COVID-19 origin hypotheses.

The analysis of the TF-IDF scores for words associated with the Natural Origin Hypothesis and the Lab Leak Hypothesis reveals distinct thematic focuses in the language used across both narratives (Figure 5). For the Natural Origin Hypothesis, the top term was wildlife, indicating that this word was both frequent and uniquely representative of this hypothesis. In contrast, the Lab Leak Hypothesis contains language suggestive of institutional processes and investigative discourse. [Supplementary 8].

**Figure 5:**
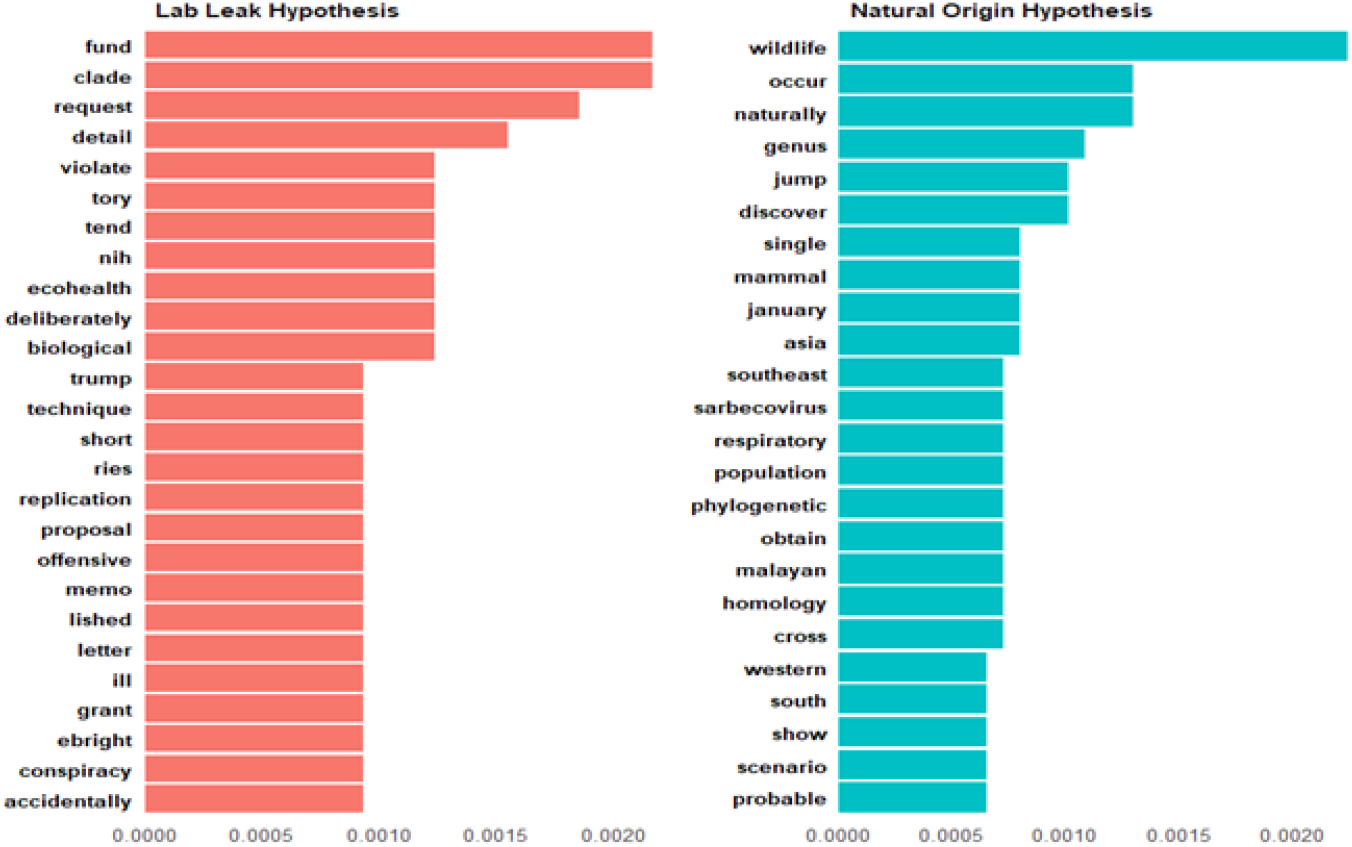
Top 20 TF-IDF ranked terms associated with the Origin hypotheses.

The keyness analysis reveals a strong lexical association between certain terms and the Natural Origin vs lab leak narrative in the COVID-19 origin debate (Figure 6). Notably, the word “bat” stands out with the highest Gš value (47.98, p < 0.0) followed by pangolin suggesting their centrality in the zoonotic transmission hypothesis. In contrast, the laboratory is associated with lab leak followed by US (P < 0.00) [Supplementary 9].

**Figure 6:**
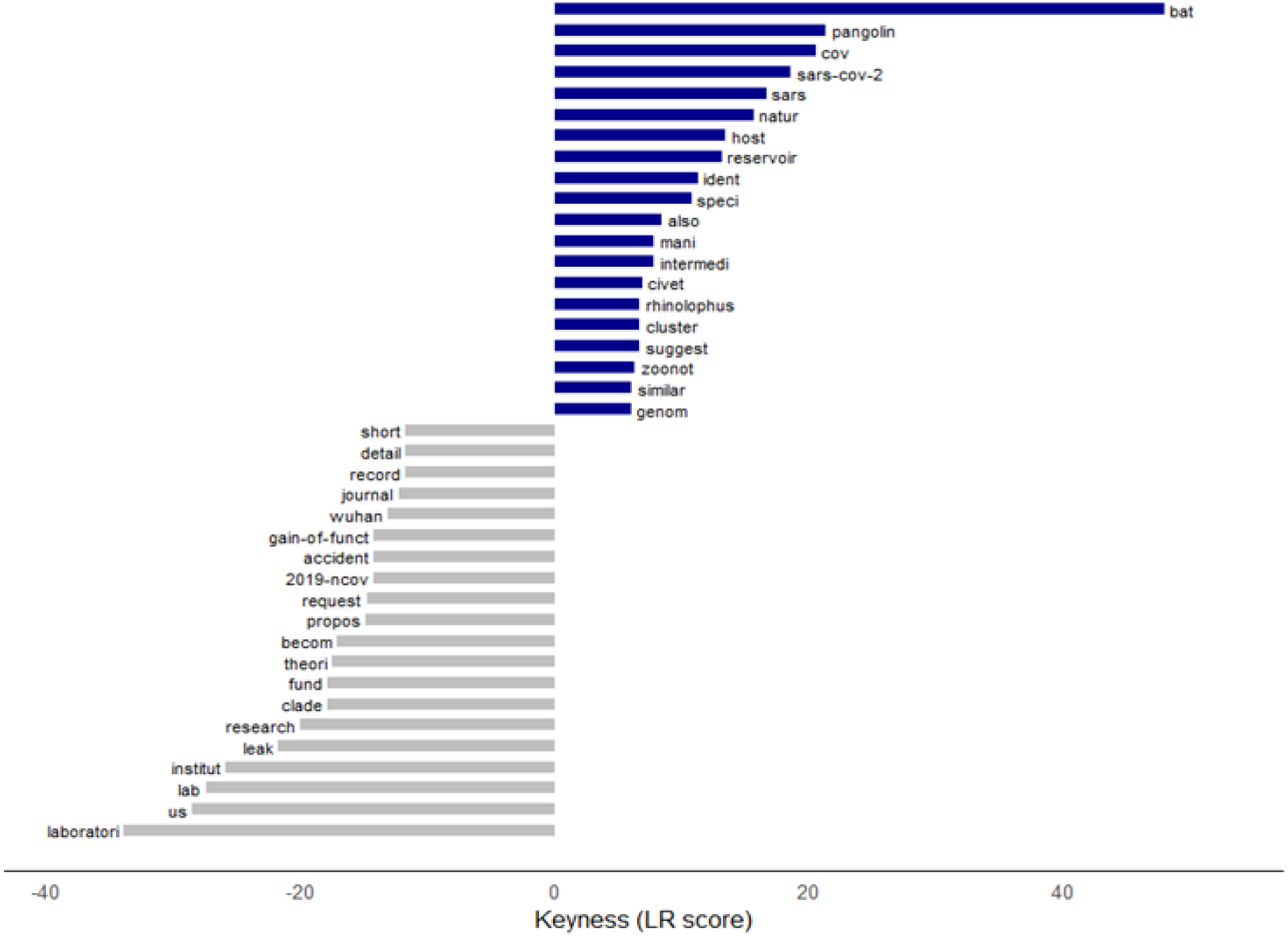
Keyness of terms associated with the Origin hypotheses

The sentiment analysis of the two narrativesLab Leak Hypothesis and Natural Origin Hypothesisreveals a notable emotional divergence in how each is discussed in the literature (Figure 7). The Natural Origin Hypothesis is associated with a positive average sentiment score of 0.398, indicating a more optimistic and affirmative language. However, the Lab Leak Hypothesis shows a negative average sentiment score of −0.124, suggesting that discourse around this theory is comparatively more negative. This indicates that languages used for this hypothesis are full of skepticism, criticism, controversy, and suspicion. The tone reflects defensive or accusatory framing, particularly in politicized or non-peer-reviewed sources.

**Figure 7:**
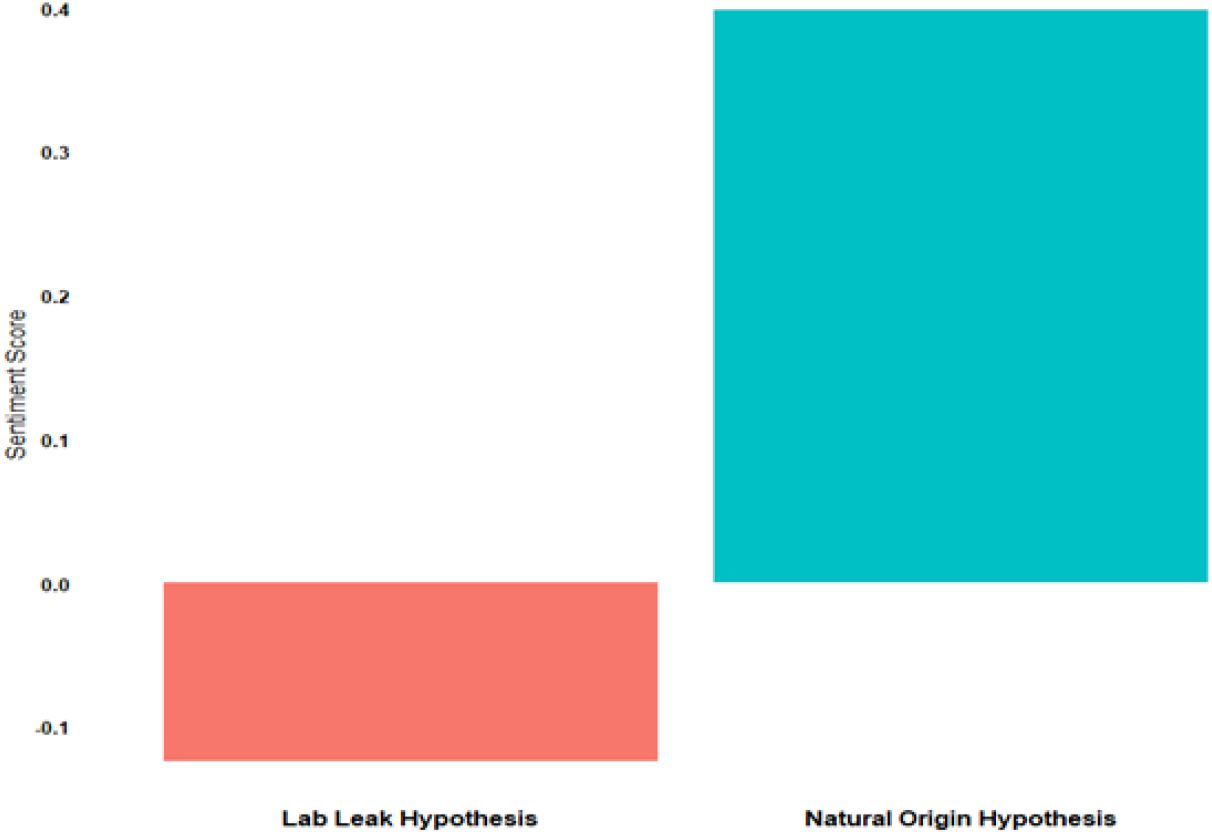
Sentiment score associated to the origin hypothesis

### Scientific Evidence supporting the origin hypothesis

A wide body of peer-reviewed genetic, virologic, and epidemiologic evidence strongly supports the hypothesis that SARS-CoV-2 emerged through natural evolutionary processes through the zoonotic spillover rather than laboratory manipulation. Several scientific statements (Segments 3, 11, 29, 263, 15, 29, 77, 178, 188, 200, 247, 155, and 357, Supplementary 10) stated that SARS-CoV-2, SARS-CoV(the virus classified as a -coronavirus), and MERS-CoV are part of the sarbecovirus lineage, a group of coronaviruses commonly found in bats, especially species within the Rhinolophus genus (Segments 186, 428, Supplementary 10) known to exist naturally and infected pangolins in Asia and Southeast Asia (Segments 2, 31, 34, 55, 56,57, 124, 228, Supplementary 10). The role of bats as reservoirs for coronaviruses, including SARS-CoV, MERS-CoV, and SARS-CoV-2, is a recurring theme across numerous research findings. Several studies demonstrated viral RNA similarities between human-infecting coronaviruses and bat viruses (Segments 68, 171, 200, Supplementary 10). Despite some hypotheses involving other animals such as snakes, genomic data have consistently ruled them out, reinforcing bats as the key reservoirs (Segment 166, Supplementary 10). It has since been established that bats host hundreds of coronavirus strains globally (Segment 6, 282, 311, 319, Supplementary 10), making them significant animal reservoirs with a broad distribution across regions like Africa, the Americas, Asia, and particularly China (Segment 8, 314, 320, Supplementary 10) which the scientists have long warned of the danger this pose to human population (Segment 1, 9, 34, Supplementary 10).

The sarbecovirus isolated from bats (Rhinolophus malayanus) in Laos was stated to exhibit high genomic similarity with SARS-CoV-2, with one particular strain, BANAL-52, showing about 96.8% similarity at the whole genome level (segment 328), while RaTG13, another bat coronavirus found in Rhinolophus affinis (horseshoe bats) in Yunnan Province, shows 96.2% similarity (segment 178, 32, 35, 46, 59, 67, 126, 139, 196, 266, 267, Supplementary 10), and that more than 780 partial coronavirus sequences have been identified in bats across 41 species infected by -coronaviruses and 31 species by -coronaviruses (Segment 14, Supplementary 10). Phylogenetic analyses and protein sequence alignments (segments 36, 80, 172, 207, 279, Supplementary 10) also support the close evolutionary relationship between SARS-CoV-2 and bat coronaviruses, and by the known role of bats as reservoirs for SARS-CoV and MERS-CoV (Segments 6, 8, 9, 34, 77, 162, 314, Supplementary 10). While some researchers have proposed possible intermediate hosts such as pangolins (segments 140, 265, Supplementary 10) and raccoon dogs (segment 304, Supplementary 10) based on genomic similarities (99% receptor-binding domain similarity in pangolin-CoVs) (Segments 138, 233, 446, Supplementary 10), the evidence indicates that SARS-CoV-2 likely originated from a recombination of bat-CoV-RaTG13-like viruses and pangolin-CoVs, possibly facilitated by environmental conditions that promote interspecies viral exchanges (segment 325, Supplementary 10). While definitive proof of pangolins as the intermediate host is lacking (segments 73, Supplementary 10), the presence of SARS-CoV-2-like viruses in smuggled pangolins from Southeast Asia (segments 379, 394, 448, 353, 226, 234, 538, Supplementary 10) and the possibility of cross-species transmission through wildlife markets or transport routes (segments 440, 452, 439, Supplementary 10) support this hypothesis. Although, these species have shown both infection and antibody responses (Segment 138, Supplementary 10). Studies also suggest that recombination events between bat and pangolin viruses may have facilitated the emergence of SARS-CoV-2 (segments 252, 514, Supplementary 10). Although genomic studies lean more heavily to-ward bats as the original host (segments 505, 538, Supplementary 10), pangolins remain a significant focus due to their high genetic similarity to the virus and their documented infection with related strains (segments 583, 265, 445, 449, Supplementary 10).

Ecological disruptions and wildlife trade have been identified as significant factors in promoting cross-species transmission (Segments 12, 41, 309, 323,326, 362,364, Supplementary 10), dense cave populations (Segments 324, 325, Supplementary 10), created an ideal condition for viral evolution and spillover. Bats, second only to rodents in mammalian diversity, are particularly adept at hosting zoonotic viruses due to traits like dense populations and high mobility (Segments 44, 316, Supplementary 10). Epidemiologically, the notion that SARS-CoV-2 originated through zoonotic spillover is strongly supported by a range of scientific literature and investigations (While direct bat-to-human transmission is theoretically possible (Segments 436, 437, Supplementary 10), with multiple segments emphasizing this route of transmission (Segments 134, 140, 226, 272, 381, Supplementary 10). Historical data support this, with evidence from the 2002-2003 SARS noting the bat-to-camel-to-human transmission route for MERS and the possible involvement of civet cats in the spillover of SARS-CoV (segment 19, 155, 357, Supplementary 10). It was stated that primates such as macaques could plausibly serve as intermediate hosts for SARS-CoV-2 due to their close genetic relationship with humans (segment 48, Supplementary 10). This aligns with the assertion in segment 52(Supplementary 10) that SARS-CoV-2 likely originated as an animal coronavirus that eventually adapted for human-to-human transmission. Strengthens this link, a connection between the first COVID-19 patients and the Wuhan wildlife market was observed (Segment 444, Supplementary 10) supported by positive environmental samples and genetic traces from early cases (Segments 97, 309, 370, 371, 375, Supplementary 10). This type of live animal market event had been recorded in the past (Segment 371, Supplementary 10) while the likelihood of direct transmission to scientists from non-bat wildlife species has been dismissed (Segment 472, Supplementary 10).

Importantly, SARS-CoV-2 lacks genetic fingerprints associated with laboratory manipulation (segments 23, 25, 107, Supplementary 10), and the furin cleavage site in SARS-CoV-2s spike protein, once cited as potential evidence of genetic engineering, is now understood to occur naturally via recombination, as it has also been identified in other coronaviruses (Segments 26, 140, 171, 250, 288, 547, Supplementary 10) supporting the idea that such insertions can emerge through recombination and natural selection (Segments 251, 289, Supplementary 10). Moreover, mutations such as N501Y are consistent with natural adaptation rather than artificial insertion enhancing the viruss transmissibility (Segment 293, Supplementary 10). SARS-CoV-2 lacks any genetic markers indicative of laboratory manipulation (Segments 23, 25, 107, Supplementary 10). While early concerns over engineered features existed, many scientists now agree that the weight of current peer-reviewed evidence points to a natural spillover event from animals to humans, likely originating in bats and possible involving intermediate hosts like pangolins or civets (Segments 78, 81, 100, 105, 144, 204, 252, 471, 473, 547, 548, 561, 104, 112, 174, 185, Supplementary 10). Although the evidence remains partly circumstantial, the majority of scientific consensus supports natural emergence as the most plausible explanation for the origin of SARS-CoV-2 (Segments 116, 191, 241, Supplementary 10). The joint WHOChina report (Segments 97, 98, 115, 300, 313, Supplementary 10) deemed a natural zoonotic spillover “likely to very likely,” while a lab-related incident was labeled “extremely unlikely.” Multiple intelligence agencies concluded that a natural zoonotic spillover was the probable origin of the virus (Segment 98, Supplementary 10). Although some questions remain, the scientific consensus overwhelmingly favors a natural emergence, rooted in bat reservoirs, with possible contributions from intermediate hosts like pangolins, raccoon dogs, or civets (Segments 100, 105, 144, 204, 252, 247, 262, Supplementary 10).

### Politicization of the COVID-19 Origin Debate

Early in the pandemic, the Trump administration suggested that the virus (SARS-CoV-2) may have originated from a lab in Wuhan, China (Segment 5, Supplementary 11). This theory gained traction with the U.S. Department of Energy and the FBI later expressing “low” and “moderate” confidence respectively in the lab-leak hypothesis (Segment 21, Supplementary 11). In contrast, other agencies remained inconclusive, stressing the need for further evidence and cooperation from China (Segment 22, Supplementary 11). Tensions escalated further when the Chinese Ministry of Foreign Affairs alleged that U.S. military personnel might have introduced the virus to Chinaa claim made without evidenceprompting reciprocal accusations from then-President Trump, who even initiated the U.S. withdrawal from the WHO (Segment 6, 9, Supplementary 11). This intense politicization led President Biden to order an intelligence review into the viruss origins in 2021, and later, in 2023, to declassify documents related to the matter (Segment 19, 20, Supplementary 11). Investigations to discover the origin had been embedded into political lines in the U.S., especially as Republican lawmakers launched numerous congressional hearings questioning figures like Anthony Fauci and agencies involved in pandemic-related research (Segment 14, 15, Supplementary 11). Meanwhile, U.S. agencies accused China of withholding data and destroying virus samples, complicating the search for answers (Segment 23, 25, Supplementary 11). China, in turn, denied these allegations, accusing the U.S. of politicizing the issue (Segment 26, Supplementary 11). The resulting geopolitical standoff was further intensified by actions such as trade sanctions on Australia after it called for an independent probe (Segment 11, Supplementary 11), and revelations that U.S. intelligence had run social media campaigns to discredit Chinese vaccines and equipment (Segment 32, Supplementary 11). Despite the WHOs efforts, access to critical data from China remained limited. It took three years for some data to be released, only to be quickly removed again from international platforms (Segment 27, 28, Supplementary 11). Experts and institutions criticized both the delay and Chinas reluctance to cooperate, underscoring a broader issue of mistrust (Segment 24, 30, Supplementary 11). Accusations also emerged regarding gain-of-function research at the Wuhan Institute of Virology (WIV), its alleged ties to the Chinese military, and the illness of WIV researchers before the pandemics official onsetfueling further speculation (Segments 16, 17, 18, Supplementary 11). The debate continues to be shaped as much by political interests and conspiracy theories as by scientific inquiry, illustrating how the pandemics origins have become a battleground for geopolitical rivalry and public accountability. Scientists have raised deep concern over the politicization of the COVID-19 pandemic highlighting a range of issues from the U.S. response to Chinese government actions this stating that discovering the origin of COVID-19 ought to be a scientific discourse (D185 and D186, Supplementary 11). The U.S. government’s accusing institutions like the CDC and FDA of flawed decision-making (Segment 74 to 105, Supplementary 11).

Interestingly, the US government report (D186, Supplementary 11) claimed that Chinese officials are said to have promoted less credible origin theories, such as transmission via frozen seafood or U.S. biolabs (segment 107, Supplementary 11). They reported that a disturbing narrative emerges of suppression and censorship: researchers like Professor Zhang, who isolated and sequenced the virus early on, were silenced and their labs shut down (segment 110, Supplementary 11). The report stated that theres evidence that Chinese authorities banned the sharing of outbreak data (segments 113, Supplementary 11) and even ordered the destruction of early virus samples (Segment 114, Supplementary 11). Crackdowns on whistleblowers (segment 115, Supplementary 11) and censorship of online discussions (segments 112, Supplementary 11) further reinforce the reports theme that the Chinese government prioritized political control over transparency. Although no proper evidence was presented to back this up.

### Global Health, Policy Implications, and Impact on the Scientific Community

The politicization of the origins of COVID-19 and past epidemics like SARS has significantly influenced global health responses, public trust, and international relations. While scientists and global health institutions tried to conduct origin-tracing, American politiciansespecially during the Trump administrationused the crisis to inflame anti-China sentiment, even voting to demand financial reparations from China and threatening to cancel U.S. debt obligations [54]. These moves, backed by significant public support, often lacked scientific grounding and ignored the complexity of zoonotic disease emergence. Such politicization fueled conspiracy theories, overshadowing legitimate investigations and increasing the risk of international conflict. This climate of blame extended to personal attacks on scientists like Dr. Anthony Fauci and misuse of scientific correspondence in attempts to fabricate controversy around COVID-19s origins. Efforts to conduct impartial inquiries, such as the Biden administrations order for intelligence agencies to investigate SARS-CoV-2s origins, were undertaken in a politically charged atmosphere, allowing misinformationoften fueled by social mediato flourish[55].

Ultimately, the politicization of disease origins, from SARS to COVID-19, has undermined global collaboration and delayed essential public health interventions. While the 2003 SARS response eventually led to the successful identification of its zoonotic origin through rapid investigation [56][57], the COVID-19 origin investigation has been significantly hindered by politicized rhetoric and blame-shifting. This underscores the need for scientific inquiry free from political interference to better prepare for and respond to future pandemics. Beliefs about the origin of COVID-19 have had profound implications for global health policy, public perception, and international relations. Unlike HIV, which required activism to gain political traction, COVID-19 is already politicized, with intelligence agencies shaping public narratives. This environment of ambiguity has left many confused and searching for scapegoats [58][59][7], resulting in the proliferation of conspiracy theories. Exposure to such conspiracy narratives has been shown to significantly affect public attitudes and behaviors, reducing support for public health measures like mask-wearing and hand hygiene [7]. These beliefs also influence policy preferences; for instance, those who believe in a lab origin are more likely to support punitive measures against China [7], while those accepting a natural origin tend to advocate for increased funding for zoonotic virus research [7]. Moreover, misinformation and competitive media framing can undermine scientific consensus and long-term public trust in science [60][61][62]. The politicization of science and the fear of persecution may deter future scientific inquiry, posing a risk not only to COVID-19 research but also to preparedness for future pandemics. Ultimately, the framing and communication of COVID-19s origins have significant downstream effects on public policy and global health strategies [63].

## RECOMMENDATIONS AND CONCLUSION

This body of evidence strongly supports a natural zoonotic origin for SARS-CoV-2. The genetic data, combined with ecological observations and epidemiological patterns, present a coherent picture of viral emergence through well-documented natural processes. While some gaps remain in our understanding of the exact transmission pathway, the overwhelming consensus from these segments points to bats as the original source, with subsequent adaptation to humans possibly facilitated by intermediate hosts in wildlife trade networks. Future pandemic prevention efforts should focus on enhanced surveillance of bat coronaviruses and stricter regulation of high-risk wildlife trade practices There is a general agreement that future coronavirus outbreaks are not just possible but likely, especially in identified hotspots such as south and southwest China. These areas have been mapped through virologic and risk studies as high-risk zones, prompting many scientists to advocate for aggressive monitoring and early warning systems for host-switching events. However, as past experiences with diseases like HIV, Ebola, Nipah, SARS, and MERS show, identifying the exact animal source or intermediate host of such pathogens often takes years. Even then, definitive conclusions are not always achievable. The challenges are compounded when investigations are hindered by political barriers, such as China’s alleged lack of transparency which some argue may prevent us from ever achieving certainty about SARS-CoV-2s origins.

Many experts underscore the need for continued, unbiased, and evidence-driven investigations, free from uninformative rhetoric. They emphasize that scientific conclusions must be based on robust and cumulative data over time. Some call for the creation of a global expert task force to conduct joint traceability studies across potentially implicated regions, while others note that public support for animal virus research is stronger among those who believe in the viruss natural origins. Further, there is a strong call for more virologic and genomic research to uncover the in vivo mechanisms of SARS-CoV-2 pathogenesis and to definitively identify intermediate hosts. This would support both the prevention of future outbreaks and the ethical oversight of high-safety laboratory research. The path to understanding COVID-19’s origins is intricate and prolonged, but global cooperation and sustained scientific inquiry are essential steps forward.

## AUTHOR CONTRIBUTIONS

Afolabi, C. O. conceived the study and led the overall design of the mixed-methods analysis. Adekunle, J. D. and Oyeniran, M. I. contributed significantly to data synthesis, data analysis, and interpretation of both the qualitative and quantitative findings. Ogu, C. K., Adegboyega, T. I., Amos, A. A., Samsudeen, O. O., Badru, K. O., Shakioye, K. O., and Alimi, A. T. supported the implementation of the inclusion and exclusion criteria and contributed to data identification and screening. Sule, H. S., Adeniyi, Y. A., and Robbert, C. O. assisted with formatting, referencing, and technical preparation of the manuscript. Oyelakin, S. O., Ayanlowo, E. J., and Alagbe, S. A. participated in reviewing the manuscript, proofreading, and correcting errors. Dr. Fagbemiro, O. provided a professional review to ensure clarity, coherence, and academic rigor.

## INFORMED CONSENT STATEMENT

This study required no informed consent statement.

## DATA AVAILABILITY

The synthesized data and all other data are available upon reasonable request.

## SUPPLEMENTARY FILES

**Supplementary 1:** Synthesized documents.

**Supplementary 2:** PRISMA Check list.

**Supplementary 3:** Source Documents for Thematic Analysis (Anonymized as D1, D2, … Dn).

**Supplementary 4:** Study-Level and Author-Level Metadata Sheets (Excel Format).

**Supplementary 5:** Thematic Categories and Supporting Narrative Data.

**Supplementary 6:** Jaccard Similarity Scores.

**Supplementary 7:** Levenshtein Distance Scores.

**Supplementary 8:** TF-IDF Weighted Keyword Tables for Competing Hypotheses.

**Supplementary 9:** Keyness Analysis Output: Term Salience in Competing Hypotheses.

**Supplementary 10:** Thematic Segments Citing Scientific Support for Origin Hypotheses.

**Supplementary 11:** Thematic Segments Highlighting Political Influences on the Origin Debate.

