## Supplementart_5 for "COVID-19 Origins: Quantifying Scientific Consensus Amid Political Polarization Through Mixed-Methods Meta-Analysis"

The textual data collected from various scientific articles, public statements, reports, and policy documents were thematically categorized to capture the breadth and complexity of the discourse surrounding the origins of COVID-19. The identified themes reflect the dominant narratives, arguments, and contextual considerations that shaped public and scientific debate. The themes are as follows:

| SN | Theme | Description |
| --- | --- | --- |
| 1 | Natural Origin Theory | Content that supports or discusses zoonotic spillover, particularly the role of bats, pangolins, and wet markets as sources of SARS-CoV-2. |
| 2 | Lab-Leak Origin Theory | Statements or arguments suggesting that the virus may have accidentally escaped from a laboratory, such as the Wuhan Institute of Virology. |
| 3 | Scientific Consensus | Contributions from the scientific community expressing consensus or broad agreement regarding the origin of the virus, often emphasizing natural spillover and dismissing unsupported lab-leak claims. |
| 4 | Politicization of the COVID-19 Origin | Texts highlighting how political actors, governments, and geopolitical tensions influenced or distorted the investigation into the virus’s origin. |
| 5 | Why Lab-Leak Fails | Evidence-based arguments and critiques explaining the limitations, inconsistencies, and lack of empirical support for the lab-leak hypothesis. |
| 6 | Prior Events | References to historical or earlier coronavirus outbreaks (e.g., SARS, MERS) and zoonotic events that contextualize the plausibility of natural origins. |
| 7 | Unknown Origin | Content that underscores uncertainties or the current lack of conclusive evidence about the origin and the intermediate host. |
| 8 | Recommendation and Future Direction | Recommendation statements given by the body of scientists. |
| 9 | Impact of Global Policy | Discussion of how international responses, policy decisions, and institutional behaviors were shaped by origin narratives, including funding, travel restrictions, and intergovernmental trust. |

:
